# Automated Aortic Regurgitation Detection and Quantification: A Deep Learning Approach Using Multi-View Echocardiography

**DOI:** 10.1101/2025.03.18.25323918

**Authors:** Christina Binder, Yuki Sahashi, Hirotaka Ieki, Milos Vukadinovic, Victoria Yuan, Meenal Rawlani, Paul Cheng, David Ouyang, Robert J. Siegel

## Abstract

**Background:** Accurate evaluation of aortic regurgitation (AR) severity is necessary for early detection and chronic disease management. AR is most commonly assessed by Doppler echocardiography, however limitations remain given variable image quality and need to integrate information from multiple views. This study developed and validated a deep learning model for automated AR severity assessment from multi-view color Doppler videos.

**Methods:** We developed a video-based convolutional neural network (R2+1D) to classify AR severity using color Doppler echocardiography videos from five standard views: parasternal long-axis (PLAX), PLAX-aortic valve focus, apical three-chamber (A3C), A3C-aortic valve focus, and apical five-chamber (A5C). The model was trained on 47,638 videos from 32,396 studies (23,240 unique patients) from Cedars-Sinai Medical Center (CSMC) and externally validated on 3369 videos from 1504 studies (1493 unique patients) from Stanford Healthcare Center (SHC).

**Results:** Combining assessments from multiple views, the EchoNet-AR model achieved excellent identification of both at least moderate AR (AUC 0.95, [95% CI 0.94-0.96]) and severe AR (AUC 0.97, [95% CI 0.96 – 0.98]). This performance was consistent in the external SHC validation cohort for both at least moderate AR (AUC 0.92, [95% CI 0.88-0.96]) and severe AR (AUC 0.94, [95% CI 0.89-0.98]). Subgroup analysis showed robust model performance across varying image quality, valve morphologies, and patient demographics. Saliency map visualizations demonstrated that the model focused on the proximal flow convergence zone and vena contracta, appropriately narrowing on hemodynamically significant regions.

**Conclusion:** The EchoNet-AR model accurately classifies AR severity and synthesizes information across multiple echocardiographic views with robust generalizability in an external cohort. The model shows potential as an automated clinical decision support tool for AR assessment, however clinical interpretation remains essential, particularly in complex cases with multiple valve pathologies or altered hemodynamics.

**Graphical Abstract:** 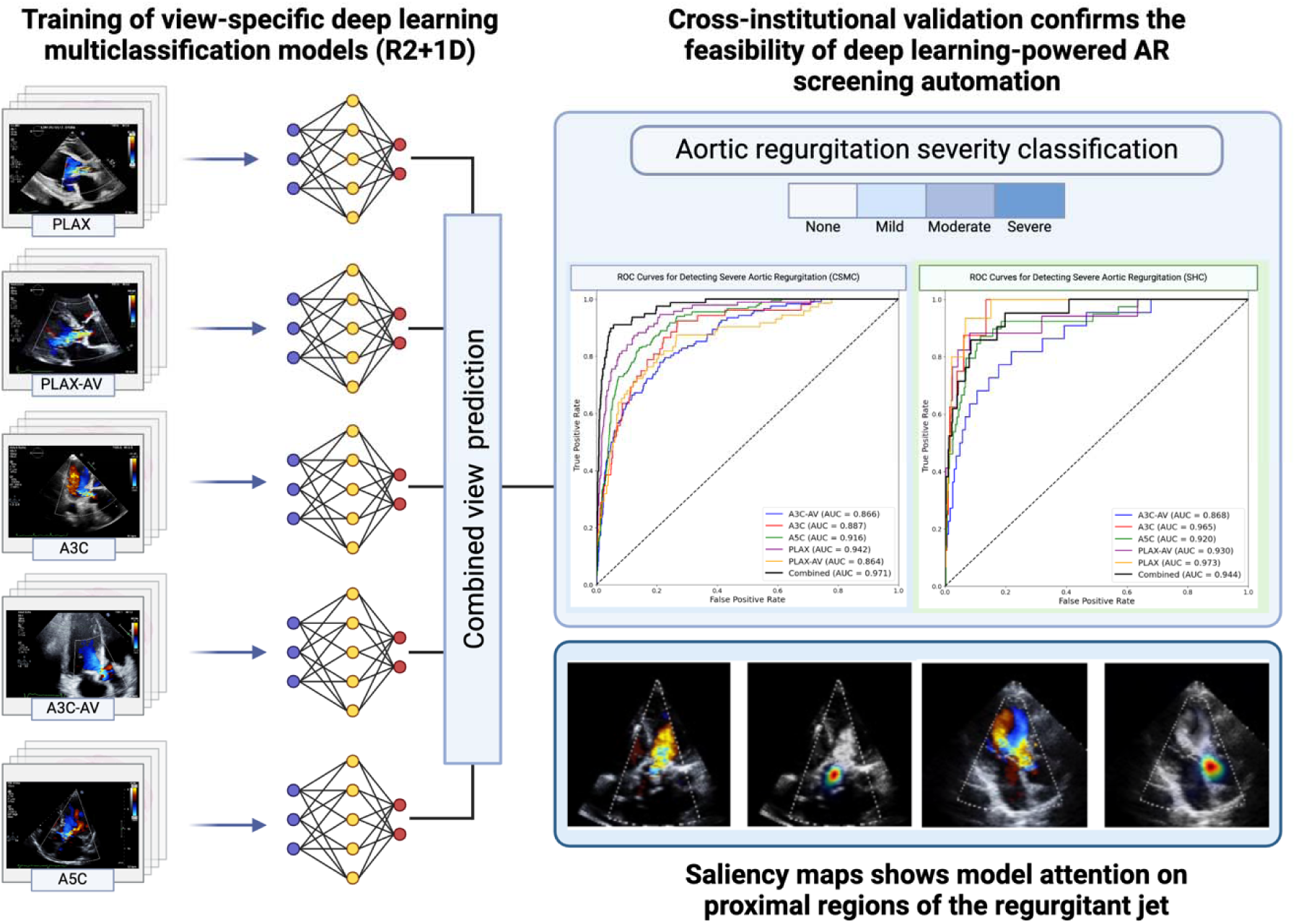

PLAX, parasternal long-axis view; PLAX-AV, parasternal long-axis view focused on the aortic valve; A3C, apical three-chamber view; A3C-AV, apical three-chamber view focused on the aortic valve; A5C, apical five-chamber view.

## Introduction

Aortic regurgitation (AR) affects patients across all age groups, with varying underlying causes. In pediatric patients and young adults, congenital aortic valve (AV) abnormalities predominate, while degenerative and rheumatic AV valve disease are the primary causes in the elderly population^1,2^. Hemodynamically significant AR triggers a cascade of left ventricular (LV) remodeling, characterized by ventricular dilatation, and progressive decline in LV function. Without intervention, this process leads to heart failure and premature death^3-6^.

Due to its cost-effectiveness and availability, Doppler echocardiography is the first-line diagnostic tool for the detection, quantification of AR-severity and assessment of hemodynamic sequelae to the LV^7^. Current guidelines recommend an integrative approach to AR grading, using a combination of qualitative, semi-quantitative and quantitative parameters^8,9^. However, some of these measurements are subjective to limitations associated with image quality or AR-jet complexity, as well as time-constraints and insufficient expertise. For these reasons, the correct identification of hemodynamically significant AR remains challenging, especially for health care providers who are not frequently confronted with the diagnosis and management of hemodynamically significant AR. Appropriate management of AR requires accurate detection and quantification of regurgitation severity to ensure the identification of the optimal timepoint for surgery as well as long term medical therapy^7,8^.

Machine learning (ML) is recently emerging as a promising tool to perform disease prediction and risk stratification, as well as disease recognition and classification. A substantial amount of ML computer vision research has been developed in the field of cardiovascular imaging, particularly echocardiography^10,11^. Even though video data adds to computational complexity as compared to still images from other diagnostic modalities, models using echocardiography video data have been shown to be able to perform tasks such as automated measurements, diagnostic classification and differential diagnosis of myocardial disease ^12-19^. Deep learning models could be a valuable tool for clinical decision making, regarding accurate assessment of disease severity and in turn, appropriate management including scheduling of follow-up intervals, and candidates for surgical intervention.

The present study aimed to develop a deep learning pipeline to quantify AR from multi-view transthoracic color Doppler echocardiography images and validate its performance in two geographically distinct cohorts. Such a high-throughput model in combination with automated LV measurements could facilitate AR quantification and support decision-making in clinical practice.

## Methods

### Study population, imaging and data source

#### CSMC Training and Evaluation Cohorts

Imaging data from a large transthoracic echocardiography (TTE) database from Cedars Sinai Medical Center (CSMC) were used for this study. Images were recorded from adult individuals receiving in-patient or out-patient care between April 2004 and June 2022 by trained and licensed sonographers on Philips EPIQ 7 or iE33. The acquisition of images followed a comprehensive and standardized protocol in-line with current recommendations^20,21^. Severity grade of AR and other parameters were extracted from echocardiography reports that had previously been evaluated and verified by board-certified cardiologists. In the clinical reports, AR severity was classified as ‘none,’ ‘trace’, ‘mild’, ‘mild to moderate’, ‘moderate’, ‘moderate to severe’ or ‘severe’. Intermediate CSMC AR severity categories were downgraded to the lower category prior to model training. Images were extracted as Digital Imaging and Communications in Medicine (DICOM) files and de-identified. Figure 1A shows an overview of the CSMC study cohort.

**Figure 1.**
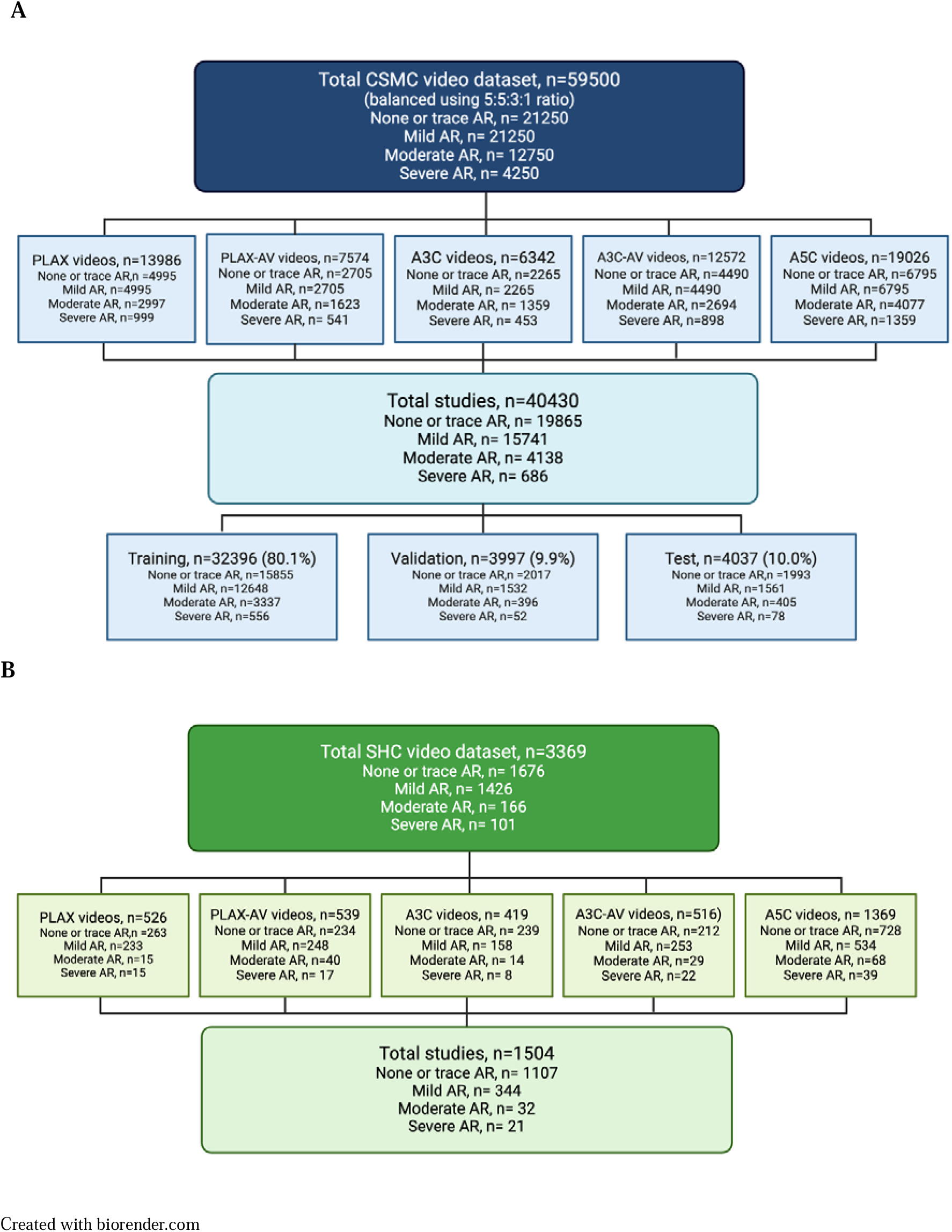
Overview of the Cedars Sinai Medical Center cohort (panel A), and the Stanford Healthcare Center cohort (panel B). TTE, transthoracic echocardiography; AR, aortic regurgitation; CD, color Doppler; PLAX, parasternal long-axis view; AV, aortic valve; A3C, apical three-chamber view and A5C, apical five-chamber view Created with biorender.com

#### SHC External Validation Cohort

A second set of TTE images from patients receiving care at Stanford Healthcare (SHC) was used for external evaluation of the trained model. The Stanford echocardiography lab uses Philips ultrasound machines and follows a similar standardized and comprehensive protocol in line with American Society of Echocardiography guidelines. AR-severity grading was extracted from clinical echocardiography reports, which had been validated by board-certified cardiologists. In the clinical echocardiography reports, AR was classified as “none”, “trace”, “mild”, “moderate”, “moderate to severe” or “severe” Figure 1B shows an overview of the SHC video validation cohort. “Moderate to severe” and “severe” were combined in the SHC cohort. This study received approval from the Institutional Review Boards of Cedars-Sinai Medical Center and Stanford Medical Center. Informed consent was waived due to the study’s use of secondary analysis of existing data.

### AI model training

We developed a deep learning classification approach for AR severity assessment using a video-based convolutional neural network. The CSMC cohort was stratified using 80% of the dataset for training, 10% for validation, and 10% as a held-out test set to evaluate model performance. To account for potential patient bias, each video was treated as an independent example, ensuring no patient overlap between the training, validation, and test cohorts. We separately trained distinct deep neural network models for videos from five different echocardiographic views (Figure 2), using the PyTorch Lightning deep learning framework. Automated view classification was pe^1^rformed as previously described^22^. We employed a video-based convolutional neural network (R2 + 1D), which has demonstrated proven effectiveness in echocardiography tasks^23^. To enhance model generalizability and prevent overfitting, we applied data augmentation techniques including random affine transformations, specifically small rotations (± 10 degrees), translations (up to 10% in x and y directions), and slight scaling (between 90-110% of original size), which introduced controlled variability to the training images. The models were trained using binary cross-entropy as the loss function with an ADAM optimizer. We initialized the networks with random weights and trained them for a maximum of 100 epochs using an initial learning rate of 1e-2 and a batch size of 24, leveraging two NVIDIA RTX 3090 GPUs. An early stopping mechanism was implemented, terminating training after 10 epochs without improvement in validation loss.

**Figure 2.**
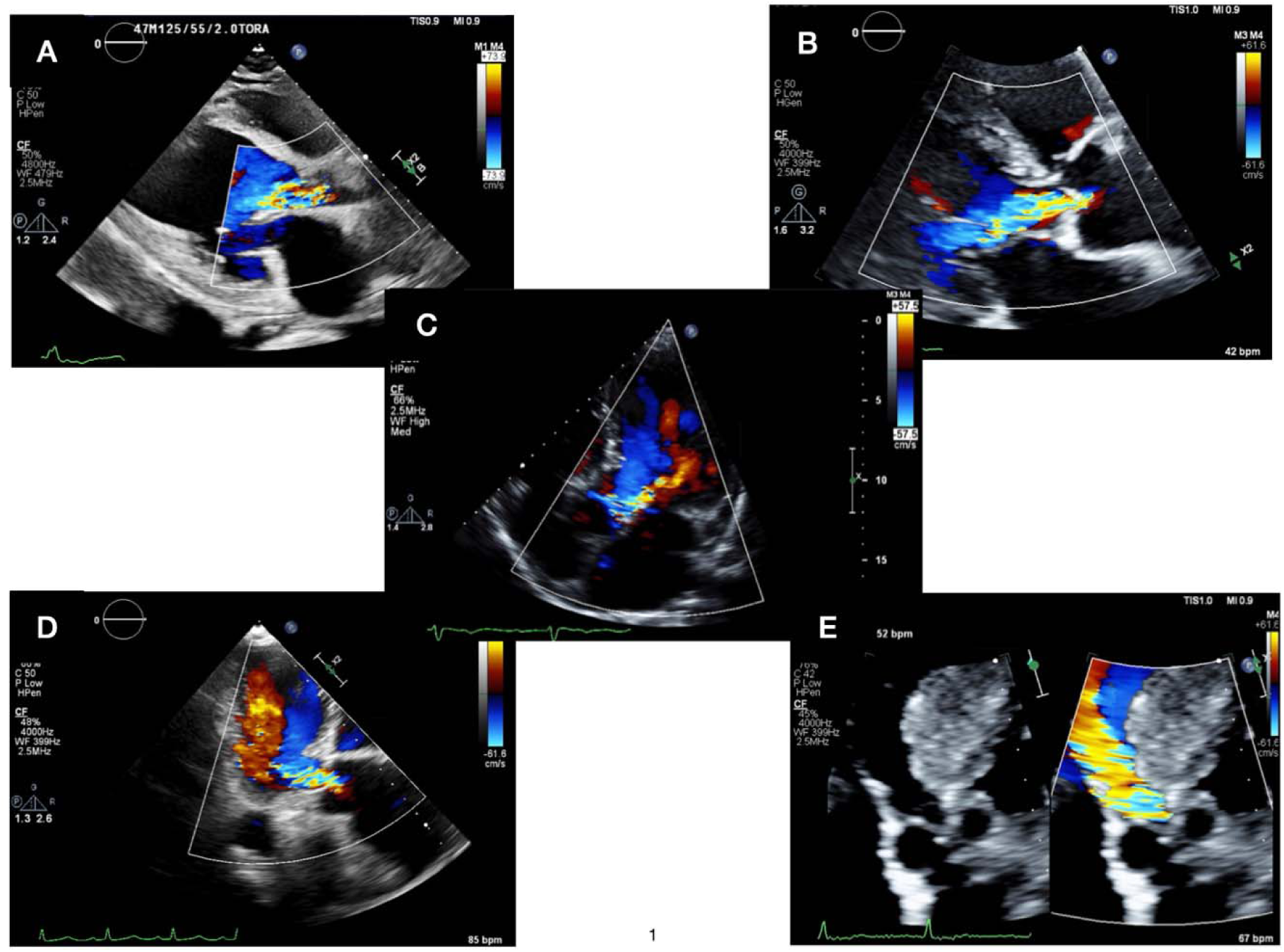
Selected Color-Doppler views for model training: parasternal long-axis view (panel A), parasternal long-axis view zoomed on aortic valve (panel B), apical five-chamber view (panel C), apical three-chamber view (panel D) and apical three-chamber view zoomed on the aortic valve (panel E).

### Statistical analysis and calculation of performance metrics

For the description of the study population, categorical variables are presented as counts and percentages, while continuous variables are expressed as median and interquartile range (IQR). Model performance was evaluated on held-out test CSMC data that was not used for model training using the weights from the epoch with the minimum loss. We calculated the area under the receiver operating characteristic curve (AUC) with 95% confidence intervals using bootstrap resampling (1000 iterations), positive predictive value (PPV), negative predictive value (NPV), recall (sensitivity), specificity and F1 score. Two classification scenarios were evaluated: detection of at least moderate AR (moderate and severe versus none/trace or mild) and detection of severe AR (severe versus all other classes). Optimal thresholds determined by calculating the Youden Index (0.772 for at least moderate AR and 0.914 for severe AR).

For study-level analysis, we combined predictions from all views within each study and selected the most severe AR class prediction to maximize sensitivity. Statistical differences between subgroups were assessed using the Kruskal-Wallis test for continuous variables and Chi-square tests for categorical variables. All analyses were performed using Python (version 3.8.0) with the scikit-learn library for machine learning metrics, scipy for statistical testing, and pandas for data management.

### Model Explainability

To understand which regions of the echocardiographic videos contributed most for AI model assessment, we employed guided backpropagation saliencey-mapping. This technique provides pixel-level attribution maps that highlight the features most influential for the model’s classification decisions. Guided backpropagation was implemented by modifying the standard backpropagation algorithm to only propagate positive gradients through ReLU activation layers, effectively suppressing negative gradients that would otherwise diminish the visualization’s clarity. We targeted the final convolutional layer (layer4) of the R(2+1)D-18 network, which captures high-level features most directly related to classification decisions. For each echocardiographic view, we processed the input video frames through the respective view-specific model. After model prediction, gradients were calculated with respect to the predicted class. The absolute values of these gradients were computed to emphasize all relevant features, followed by maximum intensity projection across the temporal dimension to create a 2D saliency map. These maps were normalized, overlaid on grayscale versions of the original frames, and smoothed with a Gaussian blur to improve visualization clarity.

## Results

### Description of the datasets

A total of 59500 videos from 40430 studies of 29081 patients were identified for model training. We enriched for cases with AR, resulting in 19865 studies (49.3 %) with ‘none or trace AR’, 15741 studies (38.9%) with ‘mild’ AR, 4138 studies (10.2%) with ‘moderate AR’ and 686 studies (1.7%) with ‘severe AR’. The model was trained on 11,258 PLAX videos, 6,054 PLAX videos focusing on the aortic valve, 5,119 A3C videos, 9,979 A3C videos focusing on the aortic valve and 15,228 A5C videos (Figure 1A). There was no significant difference in the frequency of demographic characteristics or comorbidities across AR severity groups. A detailed description of the characteristics of the cohort CSMC is shown in *Table 1 and Supplementary Table S1*. Most patients with severe AR were male (71.8%), which is in line with previous epidemiologic data describing a higher prevalence of AR in male patients^24^. More severe AR patients were more likely to have bicuspid aortic valves, larger ascending aortic diameter, larger left atrial size, and higher left ventricular ejection fractions.

**Table 1.** Characteristics of the Cedars Sinai Medical Center video cohort with selected views used for training, validation and testing.

|  | Total CSMC | Training | Validation | CSMC test | SHC |
| --- | --- | --- | --- | --- | --- |
| Videos | 59500 | 47638 (80.1) | 5814 (9.8) | 6048 (10.2) | 3369 |
| Studies | 40430 | 32396 (80.1) | 3997 (9.9) | 4037 (10.0) | 1504 |
| Patients | 29081 | 23240 (79.9) | 2889 (9.9) | 2952 (10.2) | 1741 |
| Good image quality | 58553 (98.4) | 46866 (98.4) | 5730 (98.6) | 5957 (98.5) | NA |
| Female | 25805 (43.4) | 20671 (43.4) | 2537 (43.6) | 2597 (42.9) | 1533 (46.5) |
| White | 42797 (71.9) | 34283 (72.0) | 4211 (72.4) | 4303 (71.2) | 1814 (54.9) |
| Black or African American | 6374 (10.7) | 5161 (10.8) | 609 (10.5) | 604 (10.0) | 178 (5.4) |
| Asian | 4863 (8.2) | 3843 (8.1) | 447 (7.7) | 573 (9.5) | 550 (16.6) |
| Hispanic | 6399 (10.8) | 5124 (10.8) | 698 (12.0) | 577 (9.5) | 362 (10.7) |
| Hypertension | 8168 (13.7) | 6494 (13.6) | 844 (14.5) | 830 (13.7) | 1457 (43.2) |
| Coronary artery disease | 6850 (14.4) | 6850 (14.4) | 957 (16.5) | 835 (13.8) | 1019 (30.2) |
| Atrial fibrillation | 2344 (3.9) | 1859 (3.9) | 247 (4.1) | 238 (4.1) | 1138 (33.8) |
| Heart failure | 7137 (12.0) | 5624 (11.8) | 742 (12.3) | 771 (13.3) | 1564 (46.4) |
| *Left ventricular function* |  |  |  |  |  |
| Normal ($\geq 50\%$) | 44744 (75.2) | 35706 (75.0) | 4402 (75.7) | 4636 (76.7) | 2670 (79.6) |
| Mildly reduced (LVEF 41-49%) | 6067 (10.2) | 4956 (10.4) | 522 (9.0) | 589 (9.7) | 231 (6.9) |
| Moderately reduced (LVEF 30-40%) | 3342 (5.6) | 2628 (5.5) | 374 (6.4) | 340 (5.6) | 239 (7.1) |
| Severely reduced (LVEF $< 30\%$) | 5322 (8.9) | 4328 (9.1) | 514 (8.8) | 480 (7.9) | 218 (6.5) |
| *Aortic valve morphology* |  |  |  |  |  |
| Native tricuspid | 54198 (91.1) | 43460 (91.2) | 5271 (90.7) | 5467 (90.4) | 3175 (94.2) |
| Native bicuspid | 2335 (3.9) | 1883 (4.0) | 242 (4.2) | 210 (3.5) | 103 (3.1) |
| Aortic valve prosthesis | 6985 (11.7) | 5559 (11.7) | 672 (11.6) | 754 (12.5) | 91 (2.7) |
| Bio-prosthesis/ TAVR | 6532 (11.0) | 5176 (10.9) | 628 (10.8) | 728 (12.0) | 47 (1.4) |
| Mechanical prosthesis | 453 (0.8) | 383 (0.8) | 44 (0.8) | 26 (0.4) | 44 (1.3) |
| *Valvular heart disease* |  |  |  |  |  |
| *Aortic regurgitation* |  |  |  |  |  |
| None or trace | 21250 (35.7) | 16974 (35.6) | 2158 (37.1) | 2118 (35.0) | 1676 (49.7) |
| Mild | 21250 (35.7) | 17055 (35.8) | 2061 (35.5) | 2134 (35.3) | 1426 (42.3) |
| Moderate | 12750 (21.4) | 10227 (21.5) | 1248 (21.5) | 1275 (21.1) | 166 (4.9) |
| Severe | 4250 (7.1) | 3382 (7.1) | 347 (6.0) | 521 (8.6) | 101(3.0) |
| At least moderate aortic stenosis | 6212 (10.4) | 4936 (10.4) | 651 (11.2) | 625 (10.3) | 60 (1.8) |
| At least moderate mitral regurgitation | 9191 (15.5) | 7332 (15.4) | 864 (14.9) | 995 (16.5) | 357 (10.6) |
| At least moderate tricuspid regurgitation | 9690 (16.3) | 7802 (16.4) | 911 (15.7) | 977 (16.2) | 357 (10.6) |

\*surgical or TAVR

### Performance of AR severity classification in the CSMC cohort

When analyzing the performance of the model for each view separately, we found that overall, the PLAX and the A5C views performed best, with the PLAX view achieved an AUC of 0.92 for detecting at least moderate AR and an AUC of 0.94 for severe AR and the A5C view showed comparable performance with an AUC of 0.92 for both at least moderate AR and severe AR (Table S2.1). When ensembling the predictions across all views in a study, the overall performance improved, with an AUC of 0.95 (95% CI 0.94-0.96) for detecting at least moderate AR and 0.97 (95% CI 0.96-0.98) for detecting severe AR (*Figure 3A, Figure 4A and 4C, Table 2*).

**Figure 3.**
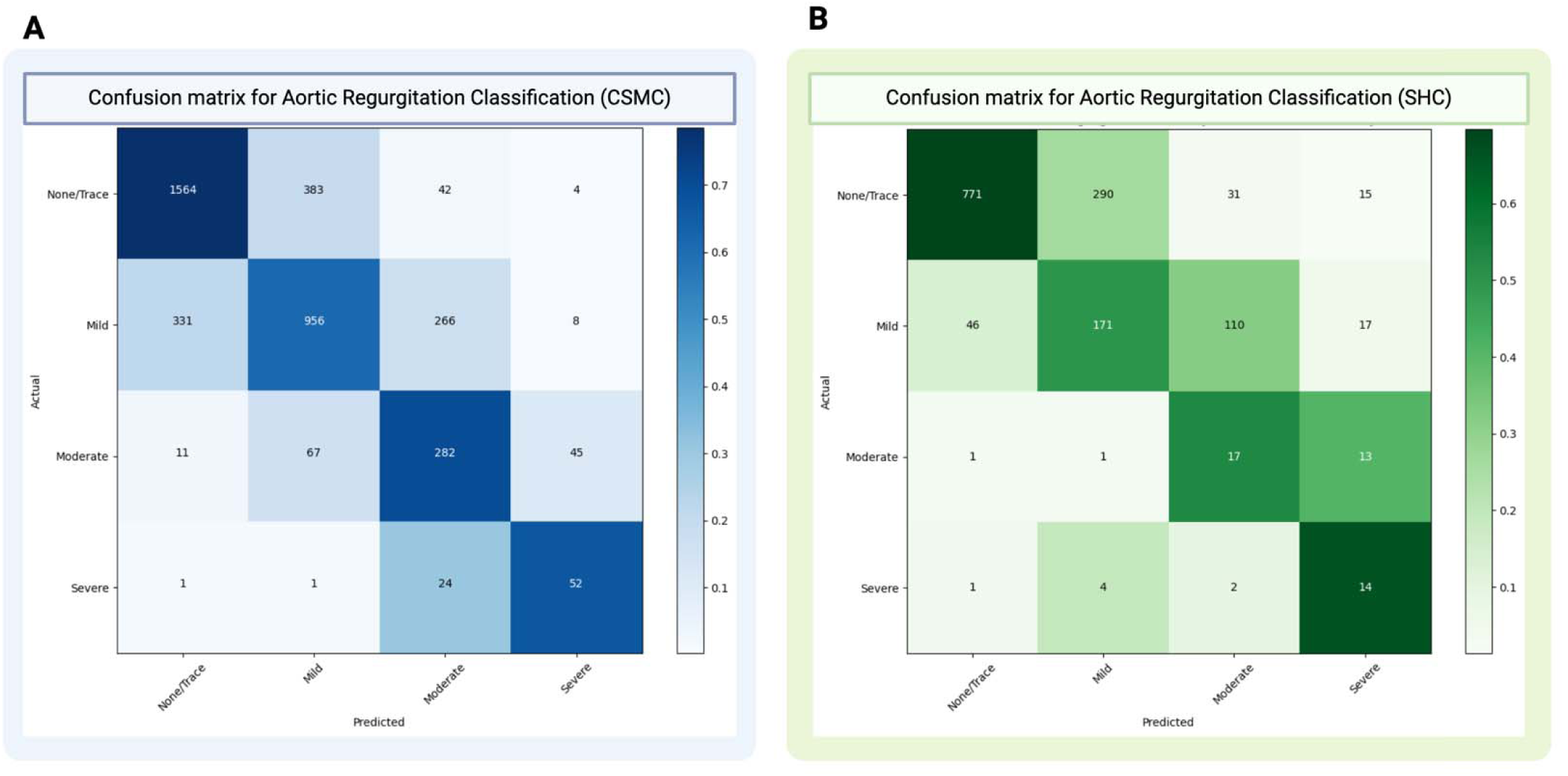
Color confusion matrix for the model’s performance to predict severity of aortic regurgitation after combining all views on a study level for the Cedars Sinai Medical Center cohort (Panel A) and the Stanford cohort (Panel B) CSMC, Cedars Sinai Medical Center; SHC, Stanford Health Care

**Table 2.** Model performance for detecting at least moderate and severe AR using a combined view approach in the Cedars Sinai Medical Center (CSMC) and Stanford Healthcare Center (SHC) cohorts.

| Site and class | AUC<br>(95% CI) | PPV<br>(95% CI) | NPV<br>(95% CI) | Sensitivity<br>(95% CI) | Specificity<br>(95% CI) | F1 score<br>(95% CI) |
| --- | --- | --- | --- | --- | --- | --- |
| CSMC |  |  |  |  |  |  |
| At least moderate AR | 0.95 (0.94-0.96) | 0.78 (0.74- 0.82) | 0.96 (0.95-0.96) | 0.67 (0.63-0.71) | 0.97 (0.97-0.98) | 0.72 (0.69-0.75) |
| Severe AR | 0.97 (0.96-0.98) | 0.80 (0.34-1.00) | 0.98 (0.98-0.99) | 0.05 (0.01-0.12) | 0.99 (0.99-1.00) | 0.10 (0.02-0.20) |
| SHC |  |  |  |  |  |  |
| At least moderate AR | 0.92 (0.88-0.96) | 0.36 (0.26-0.45) | 0.99 (0.99-1.00) | 0.74 (0.62-0.85) | 0.95 (0.94-0.96) | 0.48 (0.38-0.57) |
| Severe AR | 0.94 (0.89-0.98) | 0.99 (0.00-1.00) | 0.99 (0.98-0.99) | 0.05 (0.00-0.16) | 0.99 (0.99-1.00) | 0.09 (0.00-0.27) |

AUC, area under the receiver operating curve; 95% CI, 95% confidence interval; PPV, positive predictive value; NPV, negative predictive value; CSMC, Cedars Sinai Medical Center, SHC, Stanford Healthcare Center and AR, aortic regurgitation.

### External model validation in the SHC cohort

The model demonstrated robust generalizability in the external SHC validation cohort. When evaluating individual views, AUC values remained strong for detecting at least moderate AR across all views (0.87-0.94), comparable to the CSMC cohort performance. For severe AR detection, the PLAX view showed particularly strong discrimination in the SHC cohort (AUC 0.97). Using the combined multi-view approach, the model achieved excellent discrimination for moderate AR (AUC 0.92, [95% CI 0.88-0.96]) and severe AR (AUC 0.94, [95% CI 0.89-0.98]) in the SHC cohort (*Figure 3B, Figure 4B and C, Table 2*).

**Figure 4.**
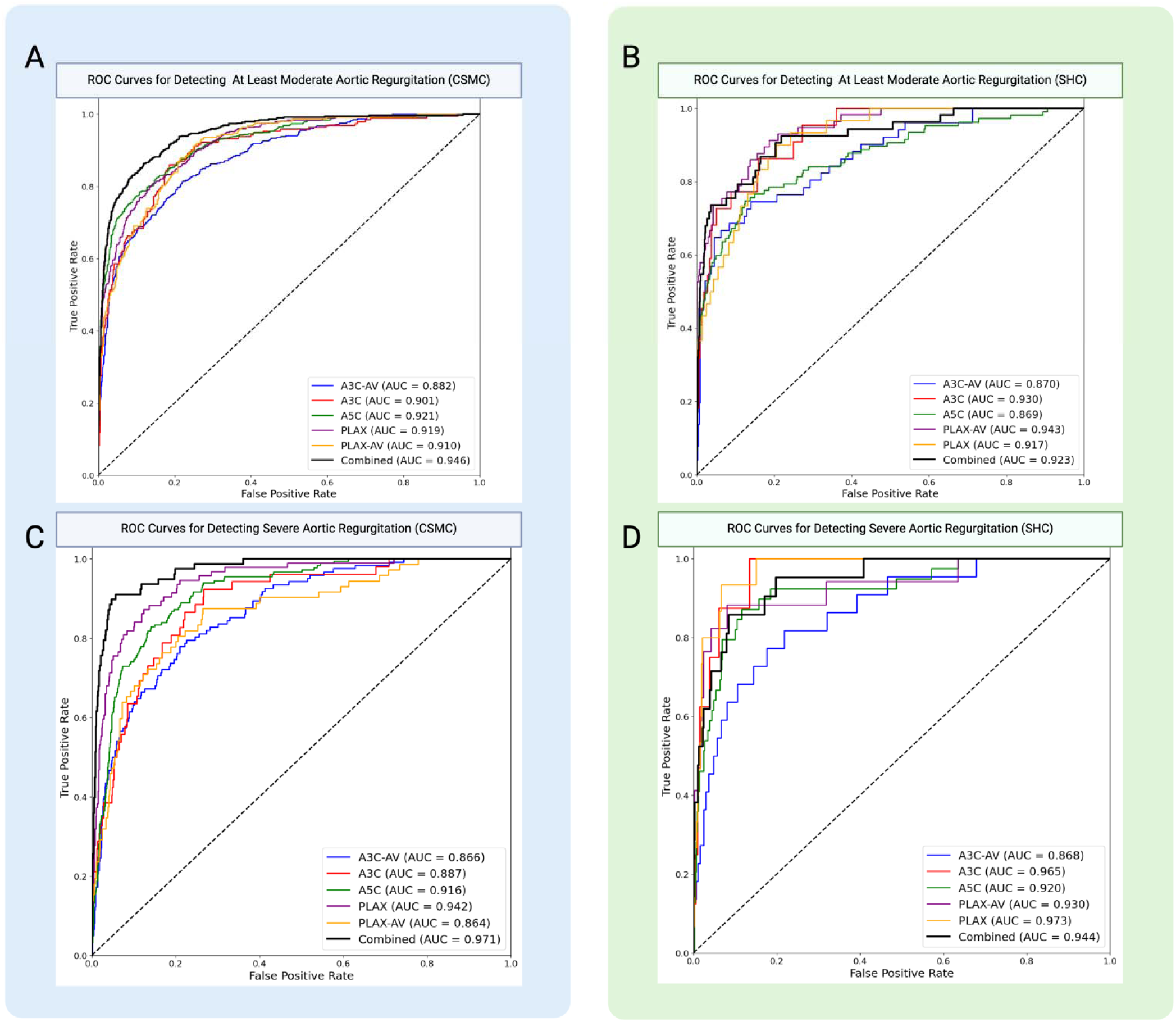
Performance of the deep learning pipeline to detect at least moderate- (Panel A and B) and severe (Panel C and D) aortic regurgitation for each view, and the combined-view model in the Cedars Sinai Healthcare Center (Panel A and C) and Stanford Healthcare Center (Panel B and D) cohort. ROC, receiver operating curve; AUC, area under the curve; CSMC, Cedars Sinai Medical Center; SHC, Stanford Health Care; A3C-AV; apical three-chamber view focused on the aortic valve; A3C, apical three-chamber view; A5C; apical five-chamber view; PLAX, parasternal long-axis view; PLAX-AV, parasternal long axis-view focused on the aortic valve.

### Subgroup analysis

The model demonstrated consistently better performance for detecting severe AR compared to at least moderate AR across most subgroups, suggesting higher discriminative ability for severe cases (Table 4). Performance was better in male patients compared to females as well as in younger patients. The model’s discriminatory ability was also better in patients with native aortic valves, whether tricuspid or bicuspid, with slightly reduced performance in videos with prosthetic aortic valves. The model maintained robust performance across varying degrees of LV size enlargement, with AUCs for detecting severe AR remaining high in patients with normal (AUC 0.96, [95% CI 0.94-0.98]), mildly dilated (AUC 0.97, [95% CI 0.94-0.99]), moderately dilated (AUC 0.95, [95% CI 0.85-0.99]) and with severely dilated LVs (AUC 0.93, [95% CI 0.85-0.99|) left ventricles. Most notably, while the model performed consistently well across normal (AUC 0.97, [95% CI 0.96-0.99]), mildly reduced (AUC 0.98, [95% CI 0.96-0.99]), and moderately reduced (AUC 0.95, [95% CI 0.88-0.99]) LV function, there was a performance drop in patients with severely reduced LV function (AUC 0.78, [95% CI 0.56-0.98]).

**Table 3.** Model performance in predefined subgroups in the Cedars Sinai Medical Center cohort.

| Subset | Model performance to detect at least moderate AR | Model performance to detect severe AR |
| --- | --- | --- |
| **Image quality** |  |  |
| Good quality | 0.94 (0.93-0.95) | 0.96 (0.94-0.98) |
| Poor quality | 0.94 (0.85-0.99) | 0.84 (0.93-0.96) |
| Female | 0.93 (0.91-0.95) | 0.95 (0.92-0.98) |
| Male | 0.95 (0.93-0.96) | 0.97 (0.94-0.99) |
| **Age** |  |  |
| ≥ 74* years | 0.91 (0.89-0.93) | 0.95 (0.90-0.98) |
| < 74* years | 0.95 (0.94-0.97) | 0.97 (0.96-0.99) |
| ≥ 50 years | 0.93 (0.91-0.94) | 0.96 (0.94-0.98) |
| < 50 years | 0.95 (0.91-0.98) | 0.97 (0.94-0.99) |
| **Aortic valve morphology** |  |  |
| Native tricuspid valve | 0.93 (0.91-0.94) | 0.97 (0.94-0.98) |
| Native bicuspid valve | 0.94 (0.90-0.99) | 0.93 (0.85-0.99) |
| Prosthetic aortic valve | 0.91 (0.86-0.95) | 0.92 (0.86-0.98) |
| **Concomitant valve disease** |  |  |
| At least moderate mitral valve regurgitation | 0.89 (0.85-0.92) | 0.93 (0.86-0.98) |
| At least moderate aortic valve stenosis | 0.88 (0.83-0.92) | 0.97 (0.93-0.99) |
| At least moderate tricuspid regurgitation | 0.86 (0.81-0.90) | 0.93 (0.84-0.99) |
| **LV size** |  |  |
| Normal | 0.93 (0.92-0.94) | 0.96 (0.94-0.98) |
| Mildly dilated | 0.94 (0.88-0.98) | 0.97 (0.94-0.99) |
| Moderately dilated | 0.91 (0.83-0.96) | 0.95 (0.85-0.99) |
| Severely dilated | 0.86 (0.76-0.95) | 0.93 (0.85-0.99) |
| **LV function** |  |  |
| Normal | 0.94 (0.92-0.95) | 0.97 (0.96-0.99) |
| Mildly reduced | 0.94 (0.91-0.96) | 0.98 (0.96-0.99) |
| Moderately reduced | 0.94 (0.90-0.97) | 0.95 (0.88-0.99) |
| Severely reduced | 0.84 (0.77-0.90) | 0.78 (0.56-0.98) |

\*Median age, Area under the curve (AUC) and 95% confidence intervals are shown.
AR, aortic regurgitation and left ventricle

### Explainability analysis

Guided backpropagation heatmaps confirmed that the model was consistently focusing on the AR jet when making its predictions (Figure 5, panel A-E). Notably, it focuses its attention on the aortic valve and the proximal portions of the AR jet. The visualizations suggest that the model understands the importance of the proximal flow convergence, as well as jet-origin and -direction.

**Figure 5.**
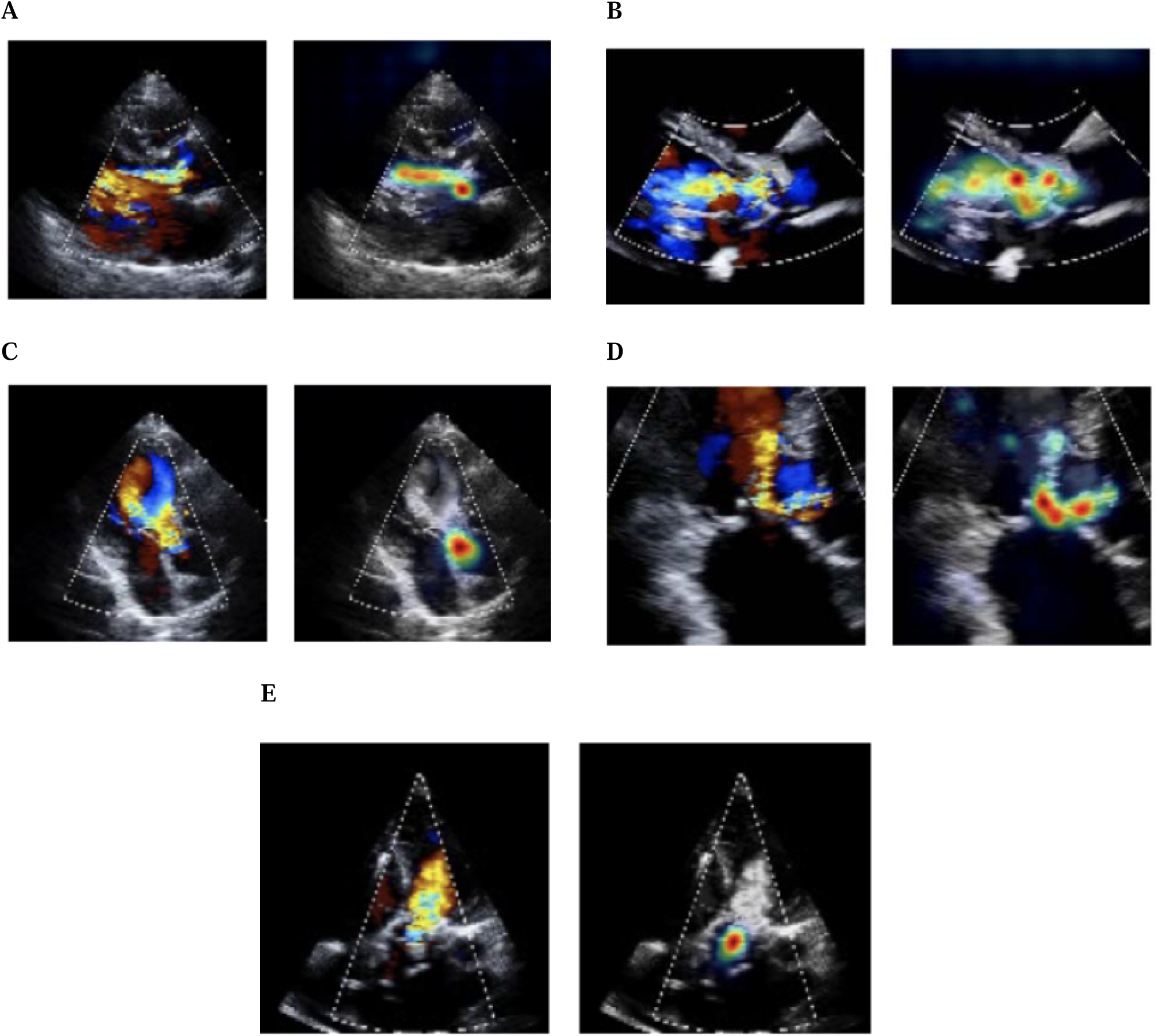
Guided backpropagation saliency map of severe aortic regurgitation cases that were predicted correctly by the model. Each panel shows the original still frame color-Doppler image (left) and the backpropagation saliency maps highlighting the regions the model focuses on for its classification prediction with the background color removed for better visualization (right). Parasternal long-axis view (panel A), parasternal long-axis view, focused on the aortic valve (panel B), apical 5 chamber view (panel C), apical 3-chamber view (panel D), apical 3-chamber view focused on the aortic valve.

### Code and Model Weights

The data set of videos and reports used to train EchoNet-AR is not publicly available because of its potentially identifiable nature; it is available for use with approval by the CSMC institutional review board. Our code and model weights are available at https://github.com/echonet/AR.

## Discussion

In this study, we developed EchoNet-AR, an automated, deep learning-based framework for the detection and severity classification of AR from color Doppler echocardiography videos. Our multi-view approach enabled accurate AR severity assessment, and the combined-view analysis integrated multiple videos to detect AR severity. Notably, these results were reproducible and robust in an external validation cohort, demonstrating the generalizability of our algorithm. EchoNet-AR showed strong performance without preselection or exclusion of comorbidities or AV morphology. While the model’s performance was exceptionally high in younger, male patients with native aortic valves, its performance dropped slightly in poor image quality, prosthetic valves and concomitant valve disease.

Consistent with clinical workflow, multiple views are necessary to accurately assess AR severity. The selection of views for our analysis was based on standard echocardiographic protocols and clinical workflow^20,25^, with views that provide complementary information about the AR jet characteristics and surrounding structures. The combined-view approach demonstrated superior performance compared to single-view analysis, aligning with human clinical practice, where multiple views are essential for accurate AR grading^25^. The guided backpropagation visualizations provide valuable insights into our model’s decision-making process for AR assessment. Images demonstrate that the model consistently focuses on hemodynamically significant regions. Particularly noteworthy is the model’s attention to proximal flow regions, specifically the flow convergence zone (PISA) and the VC area. This finding aligns with clinical practice, where these regions provide crucial information about regurgitation severity. The model’s autonomous identification of these anatomically relevant areas without explicit annotation suggests it has successfully learned clinically meaningful patterns from the training data. The consistency across different views further validates the model’s ability to recognize key diagnostic features regardless of imaging angle.

The subgroup analysis reveals robust model performance across diverse clinical scenarios, supporting its potential utility in real-world settings. Notably, the model maintained excellent discrimination (AUC ≥0.91) for detecting moderate and severe AR across various image quality conditions, patient demographics, and valve morphologies. Performance was particularly strong in younger patients (<74 years) and those with normal or mildly reduced left ventricular function. The minimal performance difference between good and poor-quality images (particularly for moderate AR detection) indicates potential resilience to common acquisition challenges encountered in everyday practice. The model’s performance on the external SHC validation cohort further demonstrated its generalizability and robustness.

## Limitations

Several limitations of our study warrant discussion. First, the ground truth for AR severity classification was based on clinical reports rather than a dedicated core lab analysis, which could introduce some variability in the reference standard. However, this approach reflects real-world clinical practice and all studies were interpreted by board-certified cardiologists following standardized guidelines. Second, while our model showed robust performance in detecting AR severity, it was not designed to perform comprehensive measurements of specific quantitative parameters such as VC width or regurgitant volume, which remain important components of clinical decision-making. Overall, the consistent performance between the CSMC and SHC datasets strengthens confidence in the model’s ability to provide reliable AR assessment in real-world clinical settings. While these results suggest the model could serve as a valuable tool for automated screening of AR, clinical interpretation and review by qualified healthcare providers remains essential to ensure appropriate patient management decisions, particularly in complex cases with multiple valve pathologies or altered hemodynamics.

## Conclusion

We developed and validated a deep learning model for accurate detection and classification of AR severity from multi-view color Doppler echocardiography videos. The model demonstrated robust performance, particularly in ruling out severe AR, and could serve as a valuable clinical decision support tool to improve the accuracy and efficiency of AR assessment in routine clinical practice.

## Acknowledgements

none

## Funding

At the time of this work, Dr. Christina Binder was a research fellow supported by a grant from the Max-Kade Foundation. Dr. David Ouyang reports support from the National Institutes of Health (R00HL157421, R01HL173487 and R01HL173526) and Alexion.

## Disclosures

Dr. David Ouyang reports consulting or honoraria for lectures from EchoIQ, Ultromics, Pfizer, InVision, the Korean Society of Echocardiography, and the Japanese Society of Echocardiography.

## Notes

### Author Declarations

Ethics committee/IRB of Cedars-Sinai Medical Center gave ethical approval for this work Ethics committee/IRB of Stanford Medical Center gave ethical approval for this work.

