## Supplemental materials for "Automated Aortic Regurgitation Detection and Quantification: A Deep Learning Approach Using Multi-View Echocardiography"

**Supplementary materials**

**Table S1** – Characteristics of the entire Cedars Sinai Medical Center cohort on a study level according to severity of aortic regurgitation.

|  | **Total** (n=40430) | **None or trace** (n=19865) | **Mild** (n=15741) | **Moderate** (n=4138) | **Severe** (n=686) | **p-value** |
| --- | --- | --- | --- | --- | --- | --- |
| Number of videos per study | 1.0 (1.0-1.0) | 1.0 (1.0-1.0) | 1.0 (1.0-2.0) | 3.0 (2.0-4.0) | 6.0 (4.0-8.0) | <0.001 |
| Good image quality | 39674 (98.1) | 19450 (97.9) | 15492 (98.1) | 4051 (97.9) | 681 (99.3) | 0.004 |
| ***Demographics*** | | | | | | |
| Age, years | 73.0 (62.0-83.0) | 69.0 (57.0-80.0) | 78.0 (68.0-86.0) | 76.0 (66.0-85.0) | 65.0 (50.8-76.0) | <0.001 |
| Female | 17998 (44.7) | 10944 (55.3) | 7255 (46.3) | 1725 (41.9) | 192 (28.2) | <0.001 |
| Male | 22246 (55.3) | 10944 (55.3) | 8420 (53.7) | 2394 (58.1) | 488 (71.8) | <0.001 |
| BSA, m^2^ | 1.8 (1.7-2.0) | 1.9 (1.7-2.0) | 1.8 (1.6-2.0) | 1.8 (1.6-2.0) | 1.9 (1.7-2.1) | <0.001 |
| Race/ethnicity, self-identified | | | | | | <0.001 |
| White | 28851 (71.7) | 13901 (70.3) | 11501 (73.4) | 2975 (72.2) | 474 (69.7) |  |
| Black | 4745 (11.8) | 2703 (13.7) | 1584 (10.1) | 391 (9.5) | 67 (9.9) |  |
| Asian | 3141 (7.8) | 1353 (6.8) | 1337 (8.5) | 393 (9.5) | 58 (8.5) |  |
| Hispanic | 4499 (11.2) | 2592 (13.1) | 1465 (9.3) | 338 (8.2) | 104 (15.3) | <0.001 |
| ***Co-morbidities*** | | | | | | |
| Hypertension, | 5710 (14.1) | 2750 (13.8) | 2318 (14.7) | 554 (13.4) | 88 (12.8) | 0.033 |
| Coronary artery disease | 6034 (14.9) | 2974 (15.0) | 2385 (15.2) | 594 (14.4) | 81 (11.8) | 0.073 |
| Atrial fibrillation | 1612 (4.0) | 728 (3.7) | 708 (4.5) | 157 (3.8) | 19 (2.8) | 0.003 |
| Heart failure | 4932 (12.2) | 2378 (12.0) | 1956 (12.4) | 504 (12.2) | 94 (13.7) | 0.366 |
| ***AV-morphology*** | | | | | | <0.001 |
| Native tricuspid | 47926 (93.8) | 18806 (94.7) | 14999 (95.3) | 3650 (88.2) | 471 (68.7) |  |
| Native bicuspid | 892 (2.2) | 126 (0.6) | 360 (2.3) | 291 (7.0) | 115 (16.8) |  |
| AV-prosthesis | 1606 (4.0) | 931 (4.7) | 380 (2.4) | 195 (4.7) | 100 (14.6) |  |
| At least moderate AS | 3618 (9.0) | 751 (3.8) | 2070 (13.2) | 731 (17.7) | 66 (9.6) | <0.001 |
| At least moderate MR | 5311 (13.1) | 1722 (8.7) | 2488 (15.8) | 950 (23.0) | 151 (22.0) | <0.001 |
| At least moderate TR | 6029 (14.9) | 2160 (10.9) | 2822 (17.9) | 933 (22.6) | 114 (16.6) | <0.001 |
| ***Echo parameters*** | | | | | | |
| LVEDD, cm | 4.5 (4.0-5.1) | 4.4 (4.0-5.0) | 4.5 (4.0-5.1) | 4.8 (4.2-5.5) | 5.7 (5.1-6.3) | <0.001 |
| LVESD, cm | 3.0 (2.5-3.6) | 2.9 (2.5-3.4) | 3.0 (2.5-3.6) | 3.3 (2.8-3.9) | 39 (3.3-4.5) | <0.001 |
| LVEDV, mL | 79.9 (57.9-111.0) | 76.6 (55.9-103.0) | 79.0 (57.2-109.0) | 97.7 (69.9-134.0) | 152.5 (110.2-195.0) | <0.001 |
| LVESV, mL | 30.6 (20.5-49.7) | 28.7 (19.7-44.0) | 30.5 (20.2-50.2) | 41.0 (25.9-65.3) | 66.2 (43.2-93.2) | <0.001 |
| IVS, cm | 1.1 (1.0-1.3) | 1.1 (0.9-1.3) | 1.1 (1.0-1.3) | 1.1 (1.0-1.3) | 1.2 (1.0-1.4) | <0.001 |
| LVEF, % | 61.0 (52.0-66.0) | 62.0 (55.0-67.0) | 60.0 (50.0-66.0) | 59.0 (49.6-65.0) | 58.0 (50.0-64.0) | <0.001 |
| LV-SV, ml | 44.0 (32.0-59.0) | 43.2 (31.9-57.0) | 42.1 (31.0-57.0) | 50.0 (35.8-69.0) | 79.4 (56.9-105.0) | <0.001 |
| LA a/p diameter, cm | 3.9 (3.4-4.5) | 3.8 (3.3-4.4) | 4.0 (3.4-4.6) | 4.0 (3.5-4.6) | 4.2 (3.6-4.7) | <0.001 |
| LAVI, mL/m^2^ | 33.3 (23.1-49.8) | 30.5 (21.2-45.3) | 35.8 (24.8-52.6) | 38.0 (26.8-56.5) | 38.7 (26.8-56.8) | <0.001 |
| RA length, cm | 5.3 (4.7-5.9) | 5.2 (4.6-5.8) | 5.3 (4.8-6.0) | 5.4 (4.8-6.0) | 5.5 (5.0-6.1) | <0.001 |
| RA area, cm^2^ | 16.7 (13.4-21.0) | 16.0 (13.0-20.0) | 17.0 (13.9-21.8) | 17.7 (14.1-22.6) | 18.6 (15.3-23.3) | <0.001 |
| E/e’ | 10.2 (7.3-14.8) | 9.5 (6.9-13.9) | 10.8 (7.9-15.4) | 11.2 (7.9-16.4) | 10.3 (7.5-15.1 | <0.001 |
| RVEDD, cm | 3.6 (3.2-4.1) | 3.6 (3.1-4.1) | 3.6 (3.2-4.2) | 3.7 (3.2-4.2) | 3.8 (3.3-4.3) | <0.001 |
| TR Vmax, m/s | 2.5 (2.2-3.0) | 2.4 (2.1-2.8) | 2.6 (2.2-2.9) | 2.6 (2.2-3.0) | 2.6 (2.2-3.0) | <0.001 |
| AV Vmax, m/s | 1.9 (1.3-2.2) | 1.7 (1.2-2.0) | 1.9 (1.3-2.3) | 2.2 (1.5-2.7) | 2.4 (1.8-3.0) | <0.001 |
| Ascending aorta, cm | 3.2 (2.9-3.6) | 3.1 (2.9-3.4) | 3.2 (3.0-3.6) | 3.4 (3.0-3.8) | 3.7 (3.2-4.1) | <0.001 |

Counts are shown as numbers and percentages, continuous variables as median and interquartile range.
AR indicates aortic regurgitation; AVR, aortic valve replacement; TAVR, transcatheter aortic valve replacement; AS, aortic stenosis; MR, mitral regurgitation; LVEDD, left ventricular end-diastolic diameter; LVESD, left ventricular end-systolic diameter; LVEDV, left ventricular end-diastolic volume; LVESV, left ventricular end-systolic volume; IVS, diameter of the interventricular septum; LVEF, left ventricular ejection fraction; LV-SV, left ventricular stroke volume; LA left atrium; LAVI, left atrial volume index; RA, right atrium; RAVI, right atrial volume index; RVEDD, right ventricular end-diastolic diameter; TAPSE, tricuspid annular plane systolic excursion; TR, tricuspid regurgitation; AV Vmax, maximal velocity of forward flow through the aortic valve and AR-PHT, pressure half-time of the aortic regurgitation jet.

**Table S2.1**- Model performance to detect at least moderate and for severe aortic regurgitation for each separate view in the Cedars Sinai Medical Center cohort.

| Detection of at least moderate aortic regurgitation | | | | | |
| --- | --- | --- | --- | --- | --- |
|  | AUC | PPV | NPV | Recall | F1 Score |
| PLAX | 0.92 | 0.81 | 0.88 | 0.65 | 0.72 |
| PLAX-AV | 0.91 | 0.78 | 0.85 | 0.64 | 0.71 |
| A3C | 0.90 | 0.73 | 0.87 | 0.71 | 0.72 |
| A3C-AV | 0.88 | 0.77 | 0.86 | 0.66 | 0.71 |
| A5C | 0.92 | 0.75 | 0.91 | 0.79 | 0.77 |
| Detection of severe aortic regurgitation | | | | | |
| PLAX | 0.94 | 0.71 | 0.96 | 0.37 | 0.49 |
| PLAX-AV | 0.86 | 0.74 | 0.93 | 0.19 | 0.31 |
| A3C | 0.89 | 0.62 | 0.94 | 0.31 | 0.41 |
| A3C-AV | 0.87 | 0.58 | 0.94 | 0.35 | 0.44 |
| A5C | 0.92 | 0.73 | 0.92 | 0.15 | 0.25 |

**Table S2.2.-** Model performance for the detection of at least moderate and for severe aortic regurgitation for each separate view in the Stanford Healthcare Center cohort.

| Detection of at least moderate aortic regurgitation | | | | | |
| --- | --- | --- | --- | --- | --- |
|  | AUC | PPV | NPV | Recall | F1 Score |
| PLAX | 0.92 | 0.33 | 0.98 | 0.60 | 0.43 |
| PLAX-AV | 0.94 | 0.42 | 0.97 | 0.81 | 0.55 |
| A3C | 0.93 | 0.33 | 0.99 | 0.77 | 0.46 |
| A3C-AV | 0.87 | 0.29 | 0.97 | 0.75 | 0.42 |
| A5C | 0.87 | 0.36 | 0.97 | 0.69 | 0.47 |
| Detection of severe aortic regurgitation | | | | | |
| PLAX | 0.97 | 0.80 | 0.98 | 0.27 | 0.40 |
| PLAX-AV | 0.93 | 0.54 | 0.98 | 0.41 | 0.47 |
| A3C | 0.97 | 0.33 | 0.98 | 0.13 | 0.18 |
| A3C-AV | 0.87 | 0.31 | 0.98 | 0.50 | 0.39 |
| A5C | 0.92 | 0.55 | 0.98 | 0.28 | 0.37 |

AUC, area under the receiver operator curve; PPV, positive predictive value; NPV, negative predictive value; PLAX, parasternal long-axis view; PLAX-AV, parasternal long-axis view focused (zoomed) on the aortic valve; A3C, apical three-chamber view; A3C-AV, apical three-chamber view focused (zoomed) on the aortic valve) and A5C, apical five-chamber view.

**Table S3**- Model performance to detect severity categories of aortic regurgitation for Cedars Sinai Medical Center (CSMC) and Stanford Healthcare (SHC) cohorts when combining all views.

| Cohort | AR severity | AUC | PPV | NPV | Recall | F1-Score |
| --- | --- | --- | --- | --- | --- | --- |
| CSMC | None or trace | 0.85 | 0.78 | 0.75 | 0.72 | 0.75 |
|  | Mild | 0.79 | 0.68 | 0.76 | 0.59 | 0.63 |
|  | Moderate | 0.93 | 0.50 | 0.97 | 0.75 | 0.60 |
|  | Severe | 0.97 | 0.52 | 0.99 | 0.59 | 0.55 |
| SHC | None or trace | 0.77 | 0.85 | 0.44 | 0.70 | 0.77 |
|  | Mild | 0.82 | 0.51 | 0.91 | 0.72 | 0.59 |
|  | Moderate | 0.92 | 0.15 | 0.99 | 0.69 | 0.25 |
|  | Severe | 0.95 | 0.32 | 0.99 | 0.52 | 0.40 |

AR indicates aortic regurgitation, AUROC, area under receiver operating curve; PPV, positive predictive value; NPV, negative predictive value; CSMC, Cedars-Sinai Medical Center; SHC, Stanford Health Care

**Table S4**- Number of videos used for training, validation and testing for each view in the Cedars Sinai Medical Center Cohort

|  | Total  (n=59500) | Training  (n=47638) | Validation (n=5814) | Test  (n=6048) |
| --- | --- | --- | --- | --- |
| PLAX | 13986 | 11258 | 1333 | 1395 |
| PLAX -AV | 7574 | 6054 | 723 | 797 |
| A3C | 6342 | 5119 | 594 | 629 |
| A3C-AV | 12572 | 9979 | 1271 | 1322 |
| A5C | 19026 | 15228 | 1893 | 1905 |

PLAX indicates parasternal long axis view; AV, aortic valve; A5C, apical five chamber view and A3C apical three chamber view

**Table S5-** Number of videos for each view and each severity category in the Cedars Sinai Medical Center and the Stanford Healthcare Center datasets.

| *Cedars Sinai Medical Center (CSMC)* | | | | | |
| --- | --- | --- | --- | --- | --- |
|  | PLAX  (n=13986) | PLAX-AV  (n=7574) | A3C  (n=6342) | A3C-AV  (n=12572) | A5C  (n=19026) |
| None or trace | 4995 | 2705 | 2265 | 4490 | 6795 |
| Mild | 4995 | 2705 | 2265 | 4490 | 6795 |
| Moderate | 2997 | 1623 | 1359 | 2694 | 4077 |
| Severe | 999 | 541 | 453 | 898 | 1359 |
| *Stanford Healthcare Center (SHC)* | | | | | |
|  | PLAX  (n=526) | PLAX-AV  (n=539) | A3C  (n=419) | A3C-AV  (n=516) | A5C  (n=1369) |
| None or trace | 263 | 234 | 239 | 212 | 728 |
| Mild | 233 | 248 | 158 | 253 | 534 |
| Moderate | 15 | 40 | 14 | 29 | 68 |
| Severe | 15 | 17 | 8 | 22 | 39 |

PLAX indicates parasternal long axis view; AV, aortic valve; A5C, apical five chamber view and A3C apical three chamber view

**Figure S1-** ROC Curves showing the deep learning model’s performance to detect aortic regurgitation classes for parasternal long-axis view (PLAX, panel A), parasternal long-axis view focused on the aortic valve (PLAX-AV, panel B), apical three-chamber view (A3C, panel C), apical three-chamber view focused on the aortic valve (A3C-AV, panel D) and the apical five-chamber view (A5C, panel E).

**A	B**

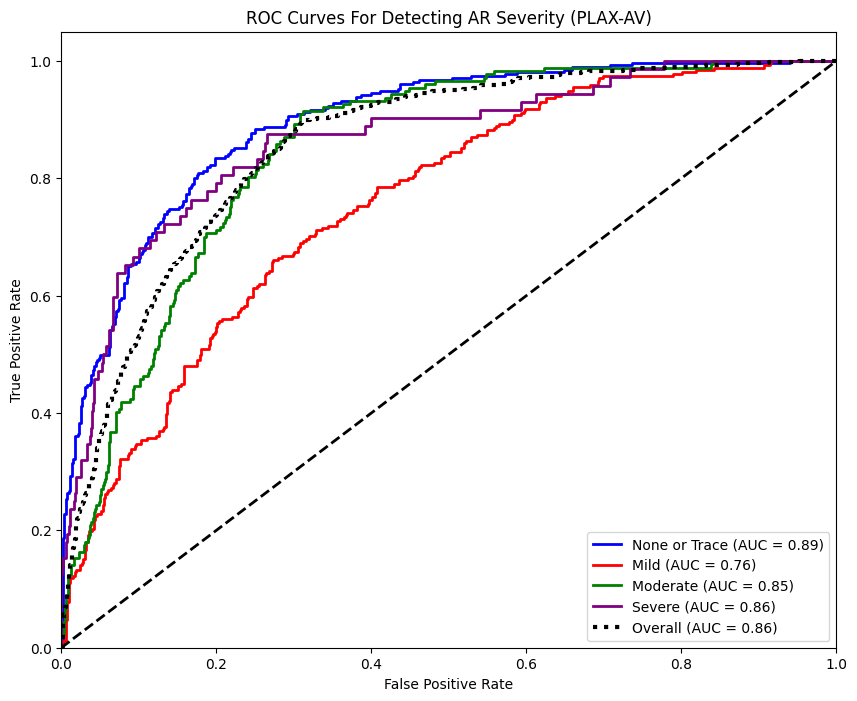

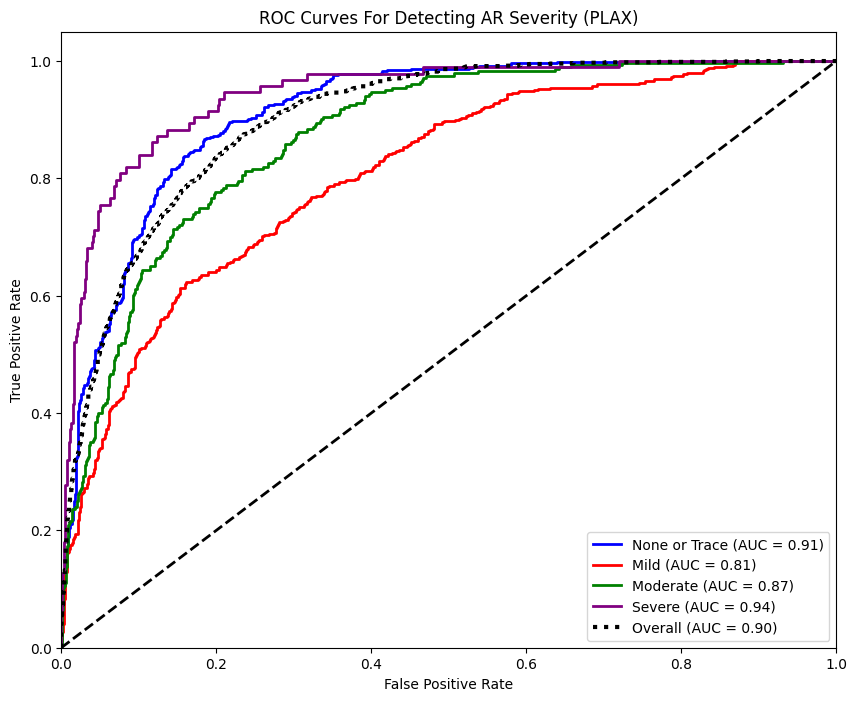

**C	D**

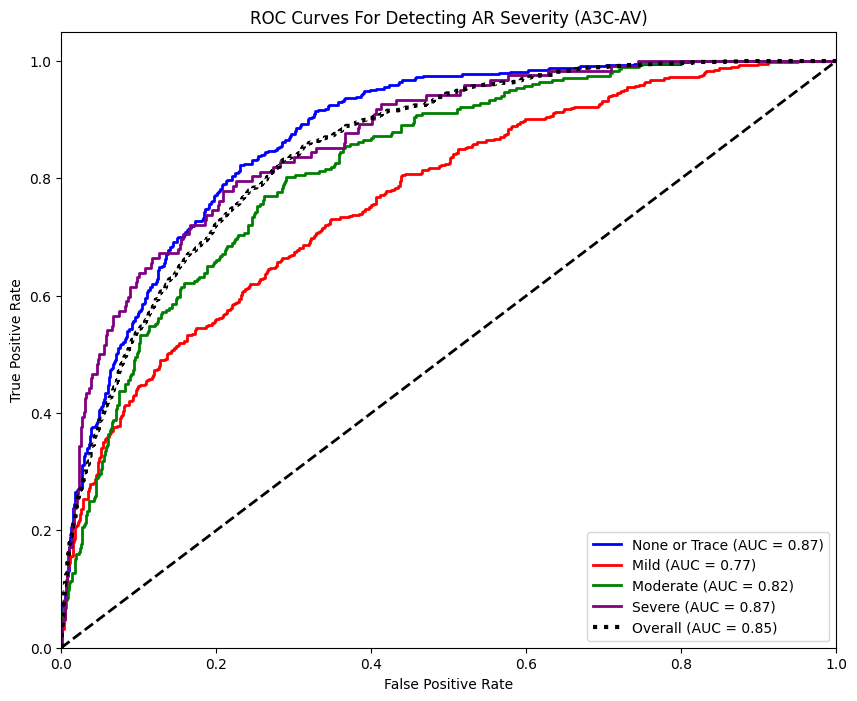

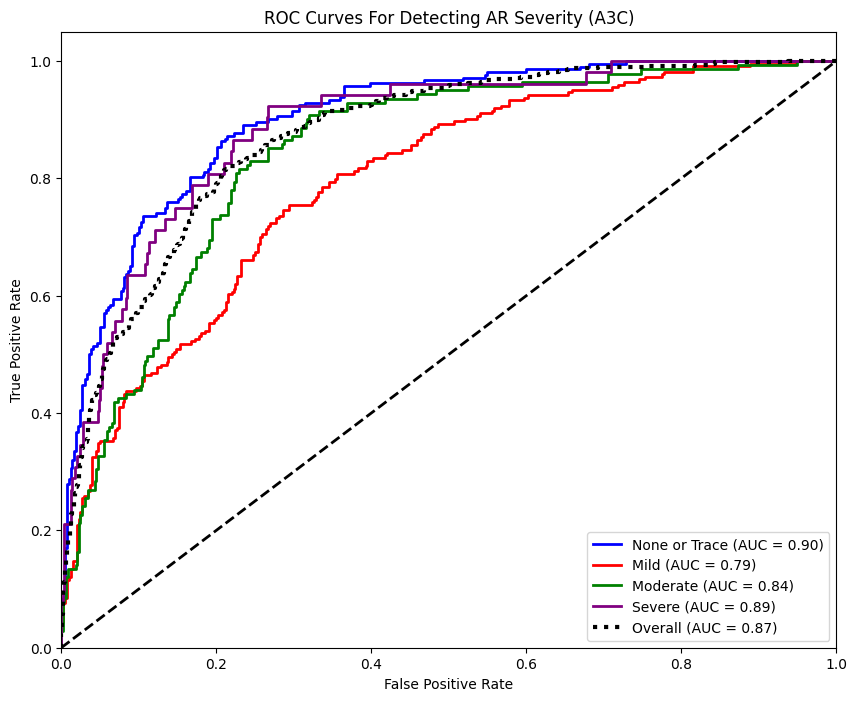

**E**

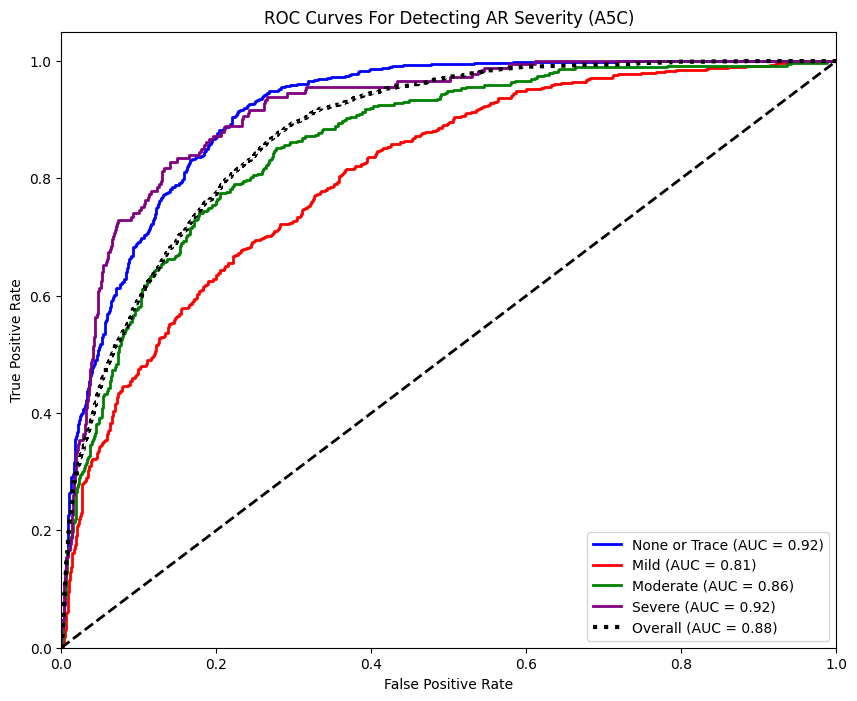
